# Deep-Learning-Based Generation of Synthetic High-Resolution MRI from Low-Resolution MRI for Use in Head and Neck Cancer Adaptive Radiotherapy

**DOI:** 10.1101/2022.06.19.22276611

**Authors:** Kareem A. Wahid, Jiaofeng Xu, Dina El-Habashy, Yomna Khamis, Moamen Abobakr, Brigid McDonald, Nicolette O’Connell, Daniel Thill, Sara Ahmed, Christina Setareh Sharafi, Kathryn Preston, Travis C Salzillo, Abdallah Mohamed, Renjie He, Nathan Cho, John Christodouleas, Clifton D. Fuller, Mohamed A. Naser

## Abstract

**Background:** Quick, low contrast resolution magnetic resonance imaging (MRI) scans are typically acquired for daily MRI-guided radiotherapy setup. However, for patients with head and neck (HN) cancer, these images are often insufficient for discriminating target volumes and organs at risk (OARs). In this study, we investigated a deep learning (DL) approach to generate high-resolution synthetic images from low-resolution images.

**Methods:** We used 108 unique HN image sets of paired 2-minute T2-weighted scans (2mMRI) and 6-minute T2-weighted scans (6mMRI). 90 image sets (∼20,000 slices) were used to train a 2-dimensional generative adversarial DL model that utilized 2mMRI as input and 6mMRI as output. Eighteen image sets were used to test model performance. Similarity metrics, including the mean squared error (MSE), structural similarity index (SSIM), and peak signal-to-noise ratio (PSNR) were calculated between normalized synthetic 6mMRI and ground-truth 6mMRI for all test cases. In addition, a previously trained OAR DL auto-segmentation model was used to segment the right parotid gland, left parotid gland, and mandible on all test case images. Dice similarity coefficients (DSC) were calculated between 2mMRI and either ground-truth 6mMRI or synthetic 6mMRI for each OAR; two one-sided t-tests were applied between the ground-truth and synthetic 6mMRI to determine equivalence. Finally, a Turing test using paired ground-truth and synthetic 6mMRI was performed using three clinician observers; the percentage of images that were correctly identified was compared to random chance using proportion equivalence tests.

**Results:** The median similarity metrics across the whole images were 0.19, 0.93, and 33.14 for MSE, SSIM, and PSNR, respectively. The median of DSCs comparing ground-truth vs. synthetic 6mMRI auto-segmented OARs were 0.84 vs. 0.83, 0.82 vs. 0.82, and 0.80 vs. 0.83 for the right parotid gland, left parotid gland, and mandible, respectively (equivalence p<0.05 for all OARs). The percent of images correctly identified was equivalent to chance (p<0.05 for all observers).

**Conclusions:** Using 2mMRI inputs, we demonstrate that DL-generated synthetic 6mMRI outputs have high similarity to ground-truth 6mMRI. Our study facilitates the clinical incorporation of synthetic MRI in MRI-guided radiotherapy.

## Introduction

Head and neck cancer (HNC) is among the most common malignancies globally (1). A core treatment modality for HNC patients is radiotherapy (RT) (2). The current clinical standard for HNC RT planning involves pre-therapy imaging using computed tomography (CT). However, adaptive RT (ART) using magnetic resonance imaging (MRI)-guided approaches offers distinct advantages over the current clinical standard, such as increased soft-tissue contrast and radiation-free intra-treatment imaging, which can be leveraged for improved tumor control and decreased side effects (3,4). Therefore, it is predicted that MRI-guided ART will play an increasingly important role in HNC patient management.

Anatomical MRI sequences, particularly T2-weighted (T2w) images, are routinely acquired during MRI-guided ART and may be used for on-the-fly segmentation of target structures, i.e., primary tumors and metastatic lymph nodes, and organs at risk (OARs) (5). Specifically, quick, low contrast resolution T2w images, typically acquired over 2 minutes, are often used for on-board setup imaging to minimize treatment times. However, these quick setup images are not always sufficient for the optimal discrimination of target structures and OARs, especially when deciding if adaptive re-planning is necessary. Longer scan times, typically performed over 6 minutes, can be employed to improve image contrast resolution, but routine use must be balanced against the patient’s comfort and the treatment schedule. Therefore, the rapid acquisition of high-resolution T2w scans is an unmet need in MRI-guided ART workflows.

Deep learning (DL) has found wide success in a variety of domains for RT-related medical imaging applications such as target and OAR segmentation (6–11) and outcome prediction (12,13). One less routinely studied domain is synthetic image generation, i.e., mapping an input image to an output image. Recent work has highlighted the utility of DL for synthetically generating CT images from MRI sequences (14–21), MRI sequences from CT images (22–26), and MRI sequences from other MRI sequences (27–31). However, to date, no studies have investigated the feasibility of using DL to generate high-resolution synthetic MRI sequences from low-resolution MRI sequences to decrease the required scan time for HNC-related imaging.

In this study, we evaluated the feasibility of generating synthetic 6-minute T2w MRI sequences from 2-minute T2w MRI sequences for use in MRI-guided ART workflows. Using paired 2-minute and 6-minute scans, we trained a DL network to generate high-quality synthetic 6-minute scans. We employed various quantitative and qualitative evaluation techniques, including a clinician-based Turing test, to demonstrate the acceptability of synthetic image generation for use in MRI-guided ART workflows. An overview of the study is shown in **Figure 1**.

**Figure 1.**
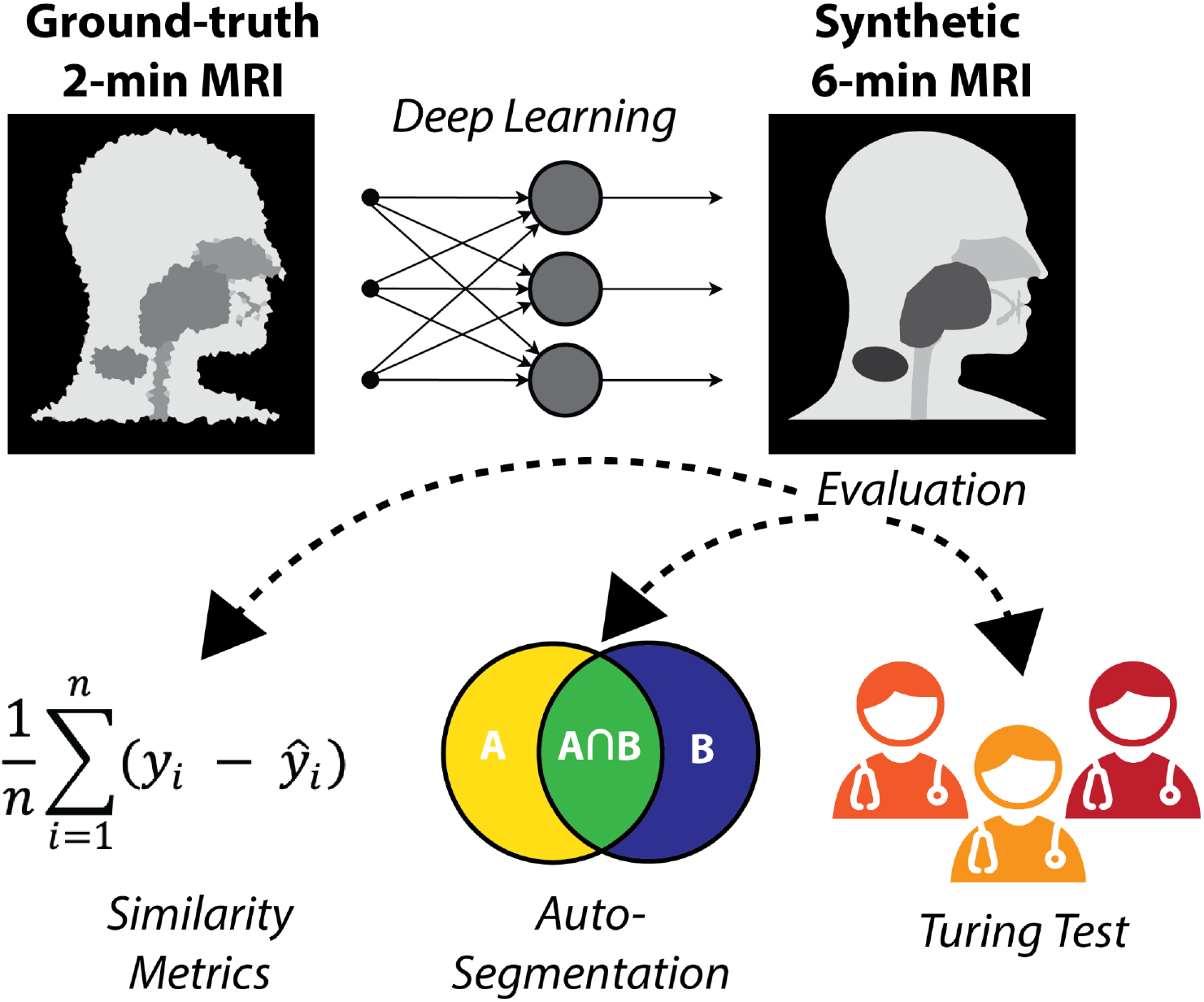
Study overview. 2-minute MRI scans are used as input to a deep learning model to generate synthetic 6-minute MRI scans. The synthetic 6-minute scans are compared to ground-truth 6-minute scans through various quantitative and qualitative methods.

## Methods

### Imaging Data

Data were retrospectively collected from a clinical trial investigating MRI-guided ART (National Clinical Trial Identifier: NCT04075305) and an internal volunteer imaging study under HIPAA-compliant protocols approved by The University of Texas MD Anderson Cancer Center’s Institutional Review Board (PA18-0341, PA14-1002, RCR03-0800). Paired 2-minute T2w MRI scans and 6-minute T2w MRI scans, acquired during the same imaging session, were collected for 53 participants (50 HNC patients; three volunteers). Details of the participants’ demographic and clinical characteristics are shown in **Appendix A**. For a subset of 10 HNC patients, multiple intra-RT paired image sets were available; the total number of paired image sets varied from 2-9 for each patient (total number of paired image sets = 56). One HNC patient had nine intra-RT 2-minute scans and one pre-RT 6-minute scan available, so a deformable image registration in ADMIRE v. 3.42 (Elekta AB, Stockholm, Sweden) was performed between the 6-minute and 2-minute scans to yield nine image sets. One volunteer had two additional paired image sets available. The remaining 21 participants (19 HNC patients pre-RT; two volunteers) had one paired image set available. In total, 108 unique paired image sets were available for use. All participants were scanned on the same scanner, a 1.5 T Elekta Unity MRI-linac device. The acquisition characteristics of the 2-minute and 6-minute scans are shown in **Table 1**. All participants were immobilized with a thermoplastic mask, which minimized differences in anatomical positioning between sequence acquisitions. All imaging data were collected in Digital Imaging and Communications in Medicine (DICOM) format.

**Table 1.** MRI sequence acquisition parameters for the 2-minute and 6-minute MRI scans used in this study.

| Acquisition Parameter | 2-minute | 6-minute |
| --- | --- | --- |
| Repetition time (ms) | 1535 | 2100 |
| Echo time (ms) | 278 | 375 |
| Echo train length | 114 | 150 |
| Flip angle (°) | 90 | 90 |
| Slice thickness (mm) | 2.0 | 2.2 |
| In-plane resolution (mm) | 0.83 | 0.68 |
| Slice gap (mm) | 1.0 | 1.1 |
| Acquisition matrix | 268x268 | 432x433 |
| Pixel bandwidth (Hz/px) | 740 | 459 |
| Number of averages | 1 | 2 |

### Data partitioning

For the purposes of model training and evaluation, we split data into separate training and test sets. The 11 HNC patients with multiple image sets, three volunteer cases, and a random sample of 21 HNC patients with single image sets were included in the training set, leading to a total of 90 unique paired image sets, i.e., ∼20,000 slices, for model training. The remaining 18 HNC patients with single image sets were used for the test set, leading to a total of 18 unique paired image sets for model testing.

### Deep Learning Network

A DL generative adversarial neural network (GAN) (32) model based on the CycleGAN architecture (33) using paired T2w 2-minute and T2w 6-minute scans was implemented in Tensorflow (34). Specifically, our model implementation draws inspiration from work performed by Johnson et al. (35), which showed promising results for image translation. The overall structure of the generative networks is based on the classic 2D Resnet encoder-decoder structure, which consists of one convolution block with a 7×7 filter and stride size of 1, two convolution blocks with 3×3 filters and stride sizes of 2, 12 residual blocks with 3×3 filters and stride sizes of 1, two corresponding up-sampling convolution blocks with 3×3 filters and stride sizes of 1, and one convolutional block with a 7×7 filter and a stride size of 1; a tanh function was used for the final prediction output. The discriminator network is a PatchGAN (36–38) containing six convolutional layers: the first four layers were composed of 4×4 filters with stride sizes of 2, while the last two layers were composed of 4×4 filters with a stride size of 1. The discriminator aims to determine whether the 142×142 image patches obtained from either generators or true images are real or fake. Similar to the original CycleGAN study (33), leaky rectified linear unit activation functions and instance normalization were used throughout the generators and discriminators. The input and output image size for the generators was 512×512 after padding, and the output patch size of the discriminators was 32×32. To obtain better quantitative synthetic results, we adopted an additional mutual information loss in addition to the original adversarial and cycle-consistency losses. The mutual information term (39) was calculated between synthetic and the ground-truth images, and the negative mutual information was minimized. The weight parameters for the adversarial loss, cycle-consistency loss, and mutual information loss were set to 1, 10, and 1, respectively. The learning rate was fixed as 2e-4 for the first half of all training epochs and was linearly decayed for the second half of training; 200 total training epochs were used. **Figure 2** shows an overview of the DL architecture components and their interaction with the loss functions.

**Figure 2.**
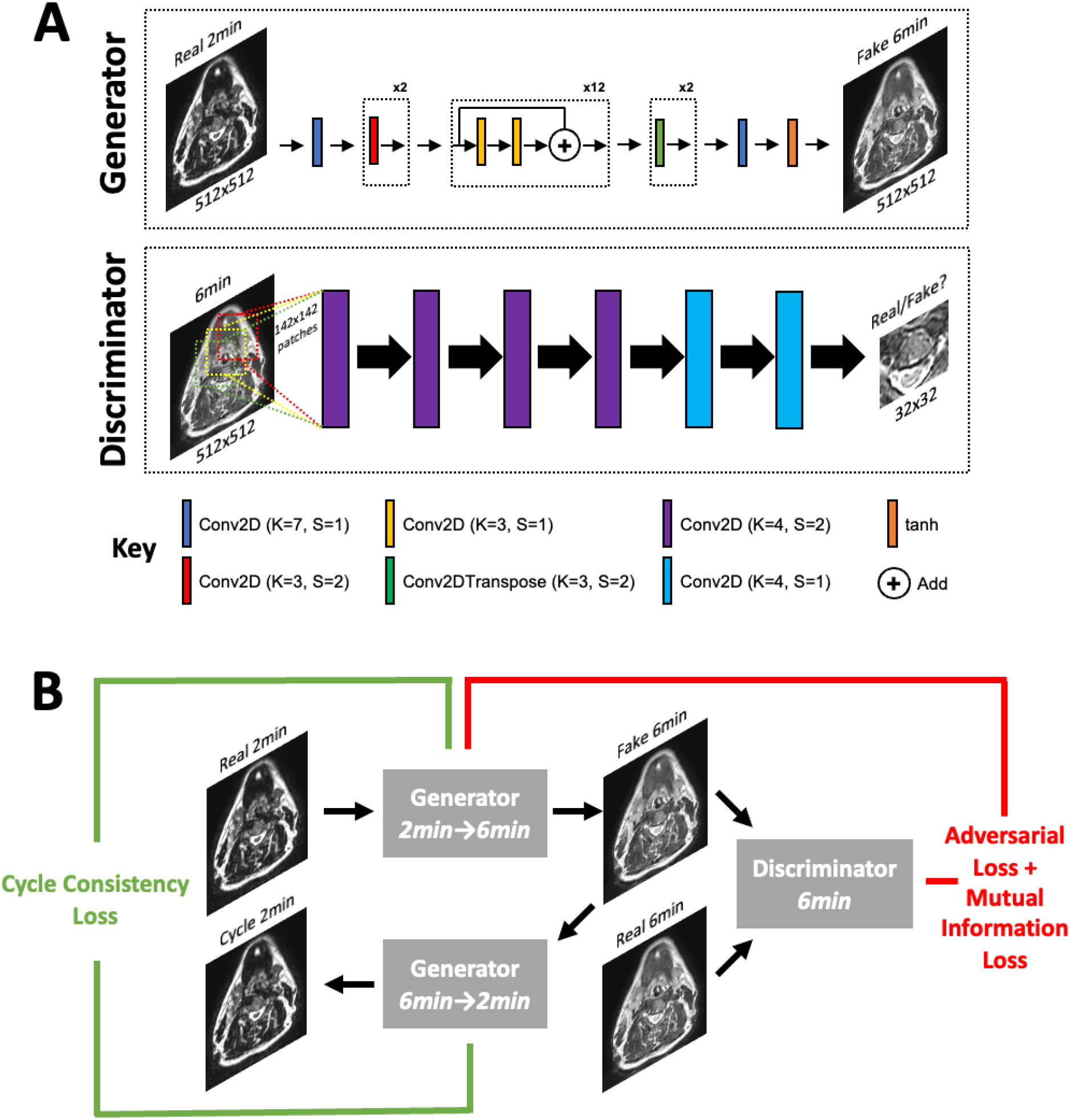
Overview of CycleGAN DL architecture and loss functions. **(A)** DL architecture. The general architecture consisted of a generator network (top) and PatchGAN discriminator network (bottom). Layers are represented as colored rectangles where K = kernel size and S = stride size. Each layer is followed by instance normalization and a leaky rectified linear unit activation function. **(B)** Loss functions used in DL network. For simplicity, only the first half of the CycleGAN is shown where a fake 6-minute scan is generated from a real 2-minute scan. An analogous process occurs to generate a fake 2-minute scan from a real 6-minute scan.

### Deep Learning Data Processing

Several simple data processing steps were performed before images were fed into the networks for training. First, 6-minute T2w images were rigidly registered with corresponding 2-minute T2w images. Previous studies have noted the importance of intensity standardization in DL-based MRI image synthesis (40); therefore, we also applied these steps to our processing pipeline. Specifically, an N4 bias field correction (41) was applied to remove additional low-frequency non-uniform artifacts. In addition, a z-score normalization, where the mean and standard deviation were calculated from the voxel values in the range of [0.25% 99.75%], was applied to 2-minute T2w images, followed by an additional rescaling to a range of [-1.0, 1.0]. The registered 6-minute T2w images were also rescaled to a range of [-1.0, 1.0]. The final trained DL model was implemented in ADMIRE v. 3.42 (Elekta AB, Stockholm, Sweden) and subsequently applied to the 2-minute test case scans to generate the synthetic 6-minute images for data analyses. Each synthetic image took approximately 1 second to generate on a V100 GPU, with an additional approximate 30 seconds after applying the N4 bias field correction. All DL inputs and outputs for the test set cases are made publicly available on Figshare: 10.6084/m9.figshare.20099252 (private until manuscript acceptance).

### Data Analyses

All analyses were performed in Python v. 3.9.7 (42). DICOM images and RT structure files were converted to Neuroimaging Informatics Technology Initiative format using the DICOMRTTool Python package (43). Subsequently, all processing operations were performed using Numpy (44) arrays. All analysis code is available on GitHub: https://github.com/kwahid/2min_6min_synthetic_MRI (private until manuscript acceptance).

### Image Similarity Evaluation

To quantitatively compare image similarity between ground-truth 6-minute scans and synthetic 6-minute scans, we used several commonly implemented metrics for image similarity. Specifically, we implemented the mean squared error (MSE), structural similarity index (SSIM), and the peak signal-to-noise ratio (PSNR). The metrics were chosen because of their widespread ubiquity in contemporary literature. All metrics were derived from the scikit-image Python package (45). SSIM was calculated using a window size of 11 as recommended in literature (46), and the PSNR was calculated using a data range based on the maximum and minimum value of the ground-truth image. Image similarity metrics were used to compare images globally (whole image) and at a region of interest (ROI) level. ROIs were based on the auto-segmented OAR structures on 2-minute scans described below (left parotid gland, right parotid gland, left submandibular gland, right submandibular gland, mandible, spinal cord, and brainstem) and an external mask of the head/neck region generated by the Otsu method (47). Bounding boxes were created around each ROI to perform the similarity calculation. Before comparison, synthetic images were resampled to the same image space as the ground-truth images using an order-3 B-spline interpolator. In addition, ground-truth images were N4 bias field corrected to ensure a fair comparison with the N4 bias field corrected synthetic images; N4 bias field post-processing was performed in ADMIRE v. 3.42 (Elekta AB, Stockholm, Sweden). Additionally, both images were z-score normalized before metric evaluation to ensure that the analysis was independent of the scale of intensity values, as suggested in previous literature (48).

### Auto-segmentation Evaluation

In **Appendix B**, we show that clinicians prefer 6-minute scans over 2-minute scans for OAR visualization. As a proxy for a human segmentation task, we implemented auto-segmentation of common HNC OARs. A previously developed DL HNC OAR auto-segmentation model available in ADMIRE v. 3.42 (Elekta AB, Stockholm, Sweden) that was trained on 2-minute T2w scans was used for the analysis. The model had previously shown superior performance to gold-standard segmentations and therefore was trusted as a reasonable proxy for clinician-generated segmentations. Additional details of the auto-segmentation model can be found in McDonald et al. (49). The left parotid gland, right parotid gland, left submandibular gland, right submandibular gland, mandible, spinal cord, and brainstem were auto-segmented for all test patients on 2-minute, ground-truth 6-minute, and synthetic 6-minute scans using the pre-trained model (representative examples shown in **Appendix C**). Dice similarity coefficient (DSC) and average surface distance (ASD) values were calculated between 2-minute scans and either ground-truth 6-minute scans or synthetic 6-minute scans for each OAR. DSC and ASD were selected because of their general ubiquity in auto-segmentation studies and ability to discriminate volumetric and surface-level segmentation quality, respectively (50). All auto-segmentation metrics were calculated using the surface-distances Python package (51). Only OARs with metric values better than previously reported interobserver variability values, as determined from McDonald et al. (49), were used for further analysis (additional details in **Appendix C**). Paired two one-sided t-tests (TOST) (52) were applied between metric values of the ground-truth 6-minute scans and synthetic 6-minute scans for each OAR to determine equivalence; p-values less than 0.05 were considered statistically significant. Equivalence bounds were determined based on the interquartile range of previous interobserver data from McDonald et al. (49) (**Appendix C, Table C1**). The Python package statsmodels (53) was used to conduct the TOST analysis.

### Turing Test

To determine whether synthetic images were distinguishable from ground-truth images by human expert observers, we implemented a Turing test inspired by Gooding et al. (54). A subset of five HNC patients with a primary diagnosis of oropharyngeal cancer with visible primary and nodal tumors on imaging were selected for the Turing test. Paired image representations were randomly generated for each slice, i.e., random allocation of images to either the left or right image. Four axial slices for six ROIs (parotid glands, submandibular glands, mandible, tumor, node) were selected for each case, leading to 100 paired images available for evaluation. Images were randomly shuffled before evaluation. Moreover, to ensure an equal comparison that is unbiased by arbitrary MRI voxel units, we applied a z-score normalization to each presented slice. Three radiation oncologists were asked to provide their best guess of which image (left or right) was real (ground-truth) and synthetic (DL-generated). In addition, clinicians were asked to denote which image they preferred overall. Finally, clinicians were asked to provide comments on why they made their decision. The test was conducted twice: first with the raw DL outputs and 2 weeks later after applying a 3×3 sharpening kernel = [[0, -0.5, 0], [-0.5, 3,-0.5], [0, -0.5, 0]] to DL outputs. To statistically evaluate the Turing test results, for each observer we implemented a two one-sided test for two proportions using an expected proportion of 0.5 with equivalence bounds of -0.3 and 0.3; the function was derived from the TOSTER R package (55) and implemented in Python through the Rpy2 Python package (56). For all statistical analyses, p-values less than 0.05 were considered statistically significant.

### Qualitative Evaluation of Failure Cases

We visually evaluated images of a select subset of cases by comparing ground-truth 6-minute scans and post-sharpened synthetic 6-minute scans. Low-, medium-, and high-performance example cases were selected on the basis of whether they were lower than, equal to, or higher than the median SSIM value of the external mask across all cases. Pixel-wise difference maps and SSIM maps between the normalized ground-truth and synthetic scans were generated to visualize uncertainties in relation to the DL synthesis process. For comparison, we also displayed the input 2-minute T2w scan for each case.

## Results

### Image Similarity Evaluation

**Table 2** shows the values of the various intensity metrics for the whole image and ROI evaluations. Two cases had surgical resections which obfuscated normal submandibular gland anatomy; therefore, these respective auto-segmented submandibular glands were not used for the ROI analysis. Generally, the whole image boasted the best median (interquartile range [IQR]) similarity metric values of 0.19 (1.13), 0.93 (0.03), and 33.14 (4.53) for MSE, SSIM, and PSNR, respectively. MSE, SSIM, and PSNR values slightly worsened when evaluated on the external mask to 0.41 (0.12), 0.81 (0.05), and 30.06 (1.32), respectively. Within the OARs, the best MSE and PSNR were for the mandible (1.16 [0.50] and 23.33 [1.83], respectively), the worst SSIM was 0.44 (0.11) for the left submandibular gland, and the worst PSNR was 18.01 (3.33) for the right submandibular gland. The brainstem simultaneously achieved the best SSIM of 0.66 (0.06) but the worst MSE of 4.99 (2.98) among the OARs. **Appendix D** shows the impact of applying various processing steps on similarity values and preliminary analyses on radiomic features.

**Table 2.**
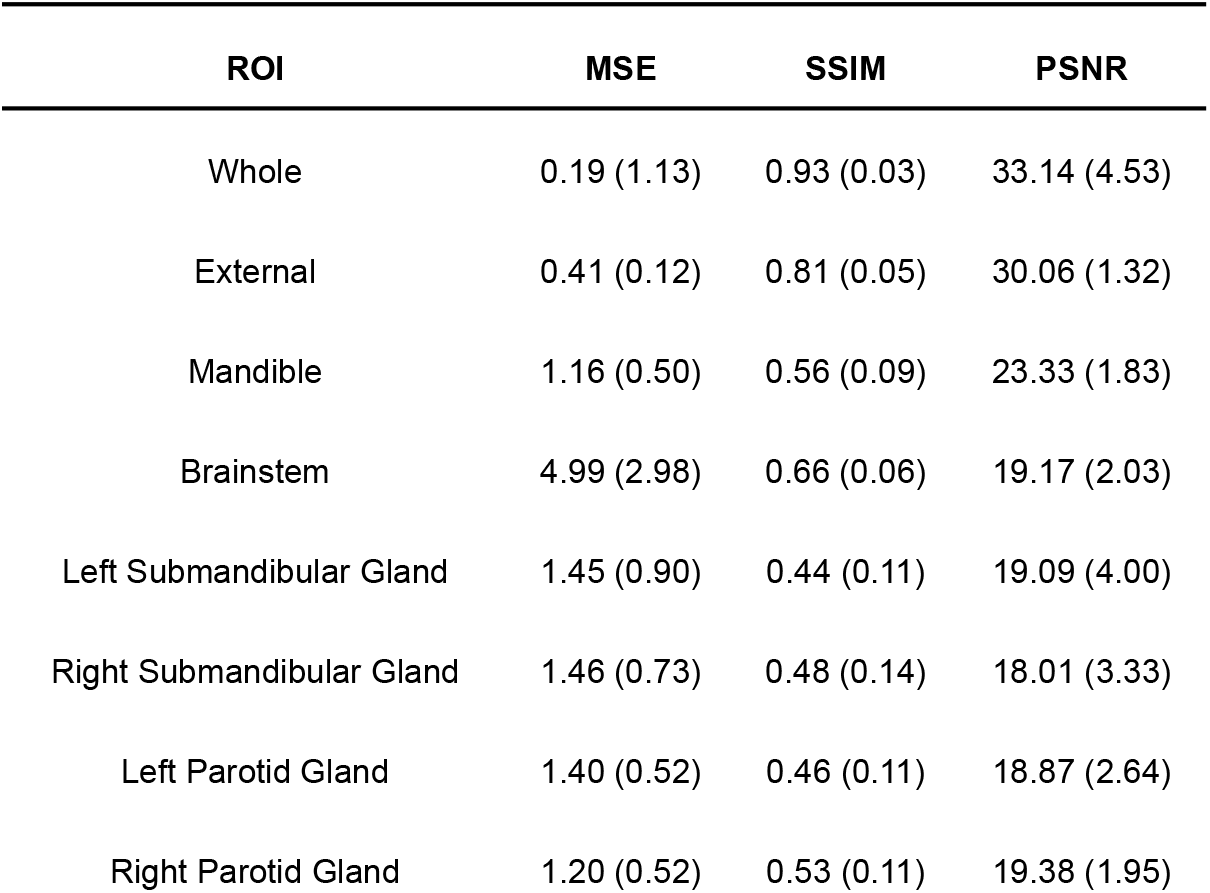
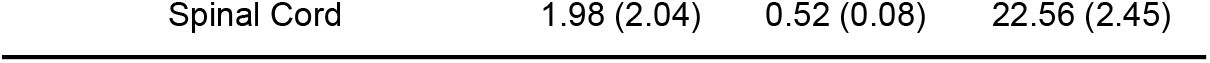
Image similarity results across the whole image and various subregions. Abbreviations: MSE = mean squared error, SSIM = structural similarity index, PSNR = peak signal-to-noise ratio.

### Auto-segmentation Evaluation

**Figure 3A** and **Figure 3B** show the distributions of the various auto-segmented OARs for the ground-truth images compared to the synthetic images for DSC and ASD, respectively. Only the right parotid gland, left parotid gland, and mandible were included in the analysis since they crossed previously recorded interobserver variability cutoffs in ground-truth images (see details in **Appendix C**). One case had a right parotidectomy; therefore, it was excluded from the right parotid analysis. The median (IQR) DSC values and equivalence test p-values comparing ground-truth vs. synthetic OARs were 0.84 (0.06) vs. 0.83 (0.06) (p=0.001), 0.82 (0.08) vs. 0.82 (0.08) (p=1.08e-7), and 0.80 (0.07) vs. 0.83 (0.07) (p=3.48e-5), for the right parotid gland, left parotid gland, and mandible, respectively. The median (IQR) ASD values and equivalence test p-values comparing ground-truth vs. synthetic OARs were 1.66 (0.87) vs. 2.15 (1.19) (p=0.048), 2.23 (1.67) vs. 2.23 (1.50) (p=4.99e-5), and 1.03 (0.61) vs. 0.80 (0.46) (p=1.57e-7) for the right parotid gland, left parotid gland, and mandible, respectively.

**Figure 3.**
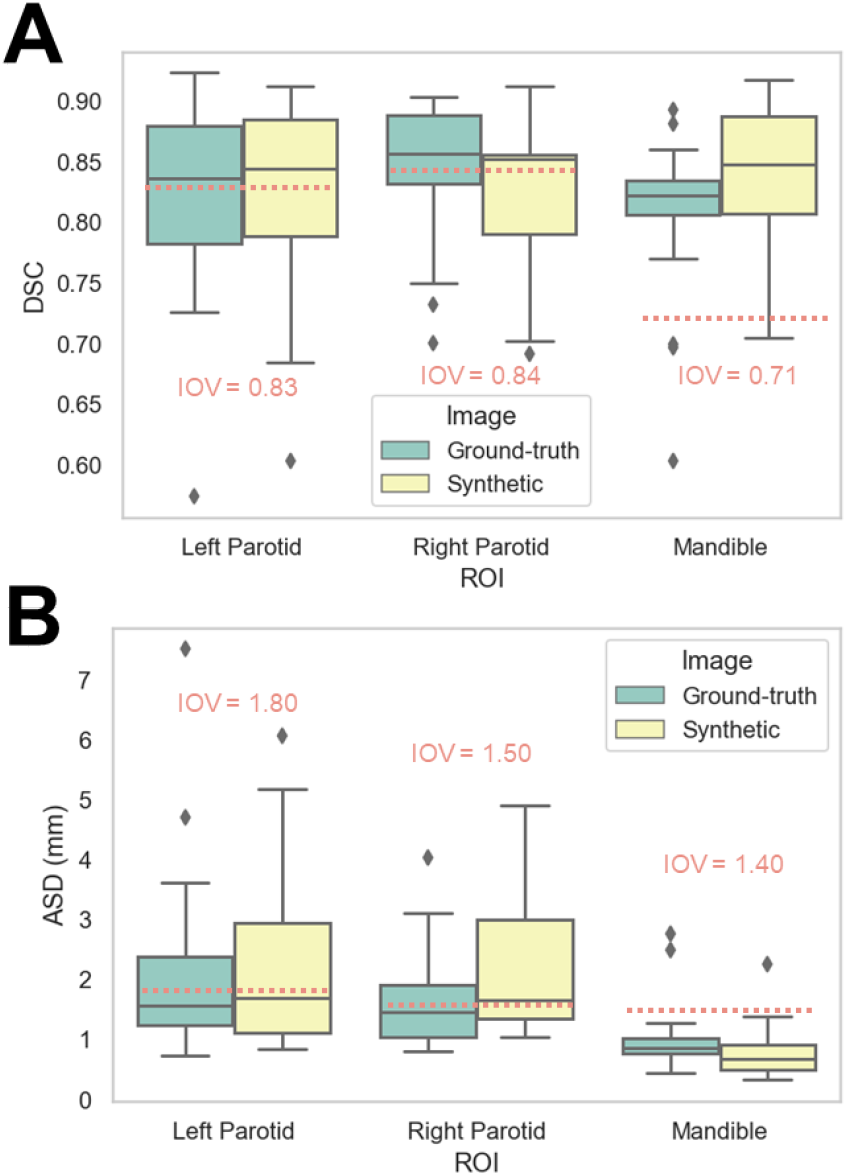
Auto-segmentation results. Auto-segmented organs at risk (left parotid gland, right parotid gland, and mandible) were generated on ground-truth (green) or synthetic (yellow) 6-minute images and compared against 2-minute images using the **(A)** Dice similarity coefficient (DSC) and **(B)** average surface distance (ASD). Red dotted lines correspond to median interobserver variability (IOV) metric values derived from clinical experts.

### Turing Test

**Table 3** shows the Turing test and clinician preference results. Significance testing for both proportions of images correctly identified and image preferences revealed equivalence between the ground-truth images and the synthetic images for all observers (p < 0.05). **Figure 4** stratifies the clinician preference results by the predominant ROI contained in each slice. All clinicians preferred the ground-truth for slices with primary tumors present, while there was no clear consensus for other regions. Observer comments for the Turing test generally focused on the ability to discriminate margins between structures (raw comments are shown in **Appendix E**). Additionally, in **Appendix E**, we show that without the application of the sharpening filter, clinicians made a clearer distinction between the synthetic and ground-truth images.

**Table 3.**
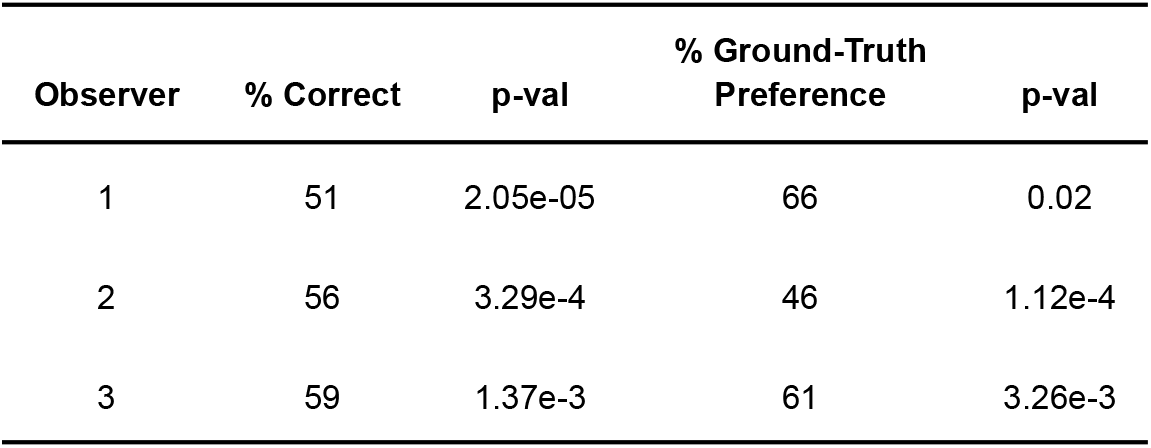
Turing test and image preference results for three physician expert observers. Each observer was asked to determine the image identity of blinded paired ground-truth or synthetic 6-minute scan slices in a randomized fashion and provide their preference. Two one-sided tests for two proportions were applied to determine whether observer estimates were equivalent to chance.

| Observer | % Correct | p-val | % Ground-Truth Preference | p-val |
| --- | --- | --- | --- | --- |
| 1 | 51 | 2.05e-05 | 66 | 0.02 |
| 2 | 56 | 3.29e-4 | 46 | 1.12e-4 |
| 3 | 59 | 1.37e-3 | 61 | 3.26e-3 |

**Figure 4.**
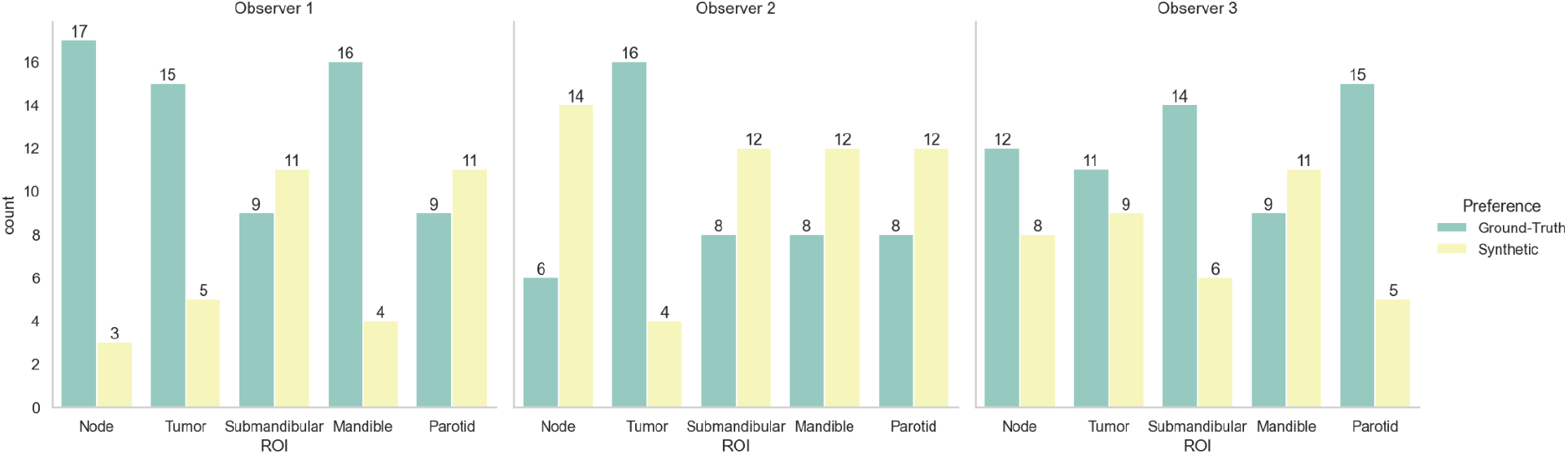
Clinician image preferences stratified by region represented in the presented image slice. Green bars correspond to ground-truth 6-minute MRI slices, while yellow bars correspond to synthetic 6-minute MRI slices.

### Qualitative Evaluation

To help visualize image similarity results, we inspected paired ground-truth and synthetic images for a few select cases based on SSIM scores of the external mask in **Figure 5**. In general, larger differences in areas of relative hyperintensity present on both 2-minute scans and ground-truth 6-minute scans were found between the ground-truth and synthetic images, e.g., fat and cerebrospinal fluid. For the low-performance case (SSIM = 0.68), which corresponds to a patient with an unknown primary tumor, a large level VI lymph node demonstrated hyperintensity on the ground-truth 6-minute scan. However, the difference map demonstrated a large discrepancy in the nodal area on the synthetic scan, likely due to a lack of visible contrast on the 2-minute scan input. An inability to synthesize the borders of the vocal folds was also noted. Relative differences were minor for most other areas of the image. For the medium-performance (SSIM = 0.81) and high-performance cases (SSIM = 0.86), which corresponded to patients with glandular and oropharyngeal primary tumors, respectively, there were no major deviations in specific regions with the exception of previously mentioned hyperintense areas. SSIM maps were generally correlated with difference maps for all cases and had lower similarity values for internal tissue structures and near tissue and background boundaries.

**Figure 5.**
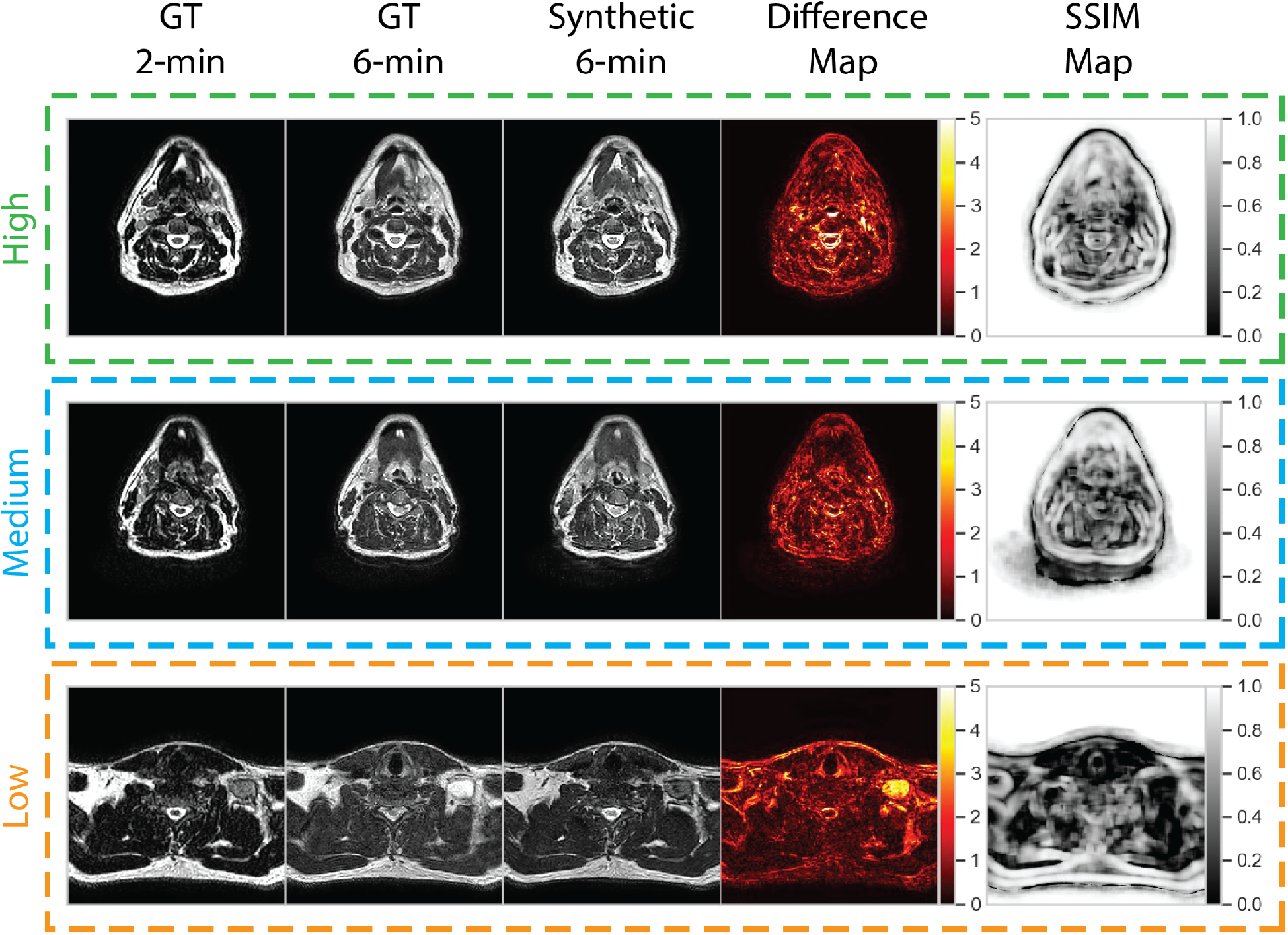
Qualitative evaluation of select cases from the test set. Cases correspond to high (green), medium (blue), and low (orange) performance relative to the median structural similarity index (SSIM) for the entire test set. Paired 2-minute scans, ground-truth 6-minute scans, synthetic 6-minute scans, absolute difference intensity maps between the ground-truth and synthetic 6-minute scans, and SSIM maps between the ground-truth and synthetic 6-minute scans are shown for each case. The high-, medium-, and low-performance cases correspond to patients with glandular (post-resection), oropharyngeal, and unknown (metastatic lymph node) primary tumors, respectively.

## Discussion

In this proof of principle study, we demonstrated the feasibility of using DL to reduce overall HNC MRI scan time by generating a 6-minute quality synthetic scan from a 2-minute scan input. A variety of evaluation techniques were performed, including similarity metric analysis, OAR auto-segmentation, and a clinician-based Turing test, which demonstrate the reasonable quality of our synthetically generated scans and potential for integration into clinical workflows. To our knowledge, this is the first study investigating the impact of DL-based synthetic image generation in the context of decreased MRI scan time in HNC and the first study to investigate synthetic image acceptability for MRI-guided ART applications.

While it is difficult to directly compare metric values between different studies due to variability in datasets and model training, our models boast similar performance to current state-of-the-art methods in related literature. Since MSE and PSNR are highly dependent on the scale of the intensity values investigated, we avoid direct comparisons to other work. However, SSIM provides an intensity scale-invariant method of calculating similarity, which allows for a surface-level comparison with other investigations. To our knowledge, the only investigation of DL-generated synthetic MRI from input MRI sequences in HNC was performed by Li et al. (31). In their study, the authors sought to generate synthetic post-contrast T1-weighted (T1w) scans from pre-contrast T1w and T2w scans, which yielded average whole-image SSIM values of 0.88 in their test set for their best-performing model. Moreover, several studies synthesizing T1w or T2w scans also reported similar SSIM values, often in the range of 0.85-0.95 (22,25,29,30). Therefore, it is encouraging that our whole-image SSIM values of 0.93 are comparable or even superior to those currently reported. It should be noted that metric values tended to worsen when evaluating specific subregions compared to the whole image. As seen in the SSIM maps of our qualitative analysis, SSIM tended to be lower for internal tissue regions. These regions may have greater intricate details that are difficult to replicate accurately. However, this trend was also noted in Li et al., where SSIM decreased from 0.86 in the whole image to 0.64 in the tumor region. Importantly, SSIM is also a function of the window size parameter, which is not currently standardized across synthetic image analysis studies. Interestingly, within the OAR regions investigated, the brainstem achieved the best SSIM but the worst MSE, indicating that the synthetic brainstem was structurally similar to the ground-truth brainstem but had a greater relative difference in raw intensity values than other OARs.

One of the main end-uses of anatomical MRIs in an ART workflow is for ROI segmentation (3). In proposed future workflows for MRI-guided ART, it is envisioned that anatomical sequences will be used to segment OARs, while functional sequences, such as diffusion-weighted and dynamic contrast-enhanced imaging, will provide greater useful information for segmenting target structures. Therefore, we have chosen to focus this study on investigating OAR-specific ROIs. Moreover, we have established physician preference for visualizing OARs on 6-minute scans compared to 2-minute scans in our supplementary analysis; therefore, it can be reasonably assumed that synthetic scans which demonstrate equivalent scan quality to a 6-minute scan would be clinically useful for OAR segmentation. While manual segmentation of OARs is still the current clinical standard, increasing effort has shown promising results for automatic OAR segmentation through DL methods (57). Our results indicate that certain auto-segmented OARs (parotid glands and mandible) are not significantly different in segmentation quality between ground-truth and synthetic scans. In essence, the previously trained auto-segmentation algorithm faced its own “artificial Turing Test” when using synthetic or ground-truth 6-minute scans as input. Subsequently, it stands to reason that a synthetic 6-minute MRI may be able to replace a real 6-minute MRI for auto-segmenting these OARs.

Unlike CT imaging, where voxel-level quantitative information can be used in the RT workflow, e.g., dose calculations, MRIs are currently mainly utilized for human-level decision making, e.g., segmentation. Arguably, the most important facet of MRI quality in current RT workflows is clinician interpretation of image quality. Therefore, the passing of a clinician-based Turing test for synthetic 6-minute scans is paramount to determining the clinical utility of this technology. In aggregate across all observers, there was no major preference given to either the ground-truth or synthetic images, indicating a similarity in the human interpretation of image quality. Moreover, observers were at most only able to correctly determine the true identity of the blinded image 59% of the time, compared to the assumed 50% if clinicians were blindly guessing. It should be noted that clinicians did have a slight preference for visualizing slices containing target volumes on ground-truth images. However, as previously noted, OAR regions are of greater interest in these anatomical sequences; therefore, whether this preference would be clinically meaningful could be debated.

Our qualitative analysis revealed that across most patients observed, there was a larger disparity in regions of relatively high intensity on ground-truth imaging, e.g., fat and cerebrospinal fluid. The larger absolute range of values may make it more difficult for the DL network to synthesize these regions precisely. Moreover, SSIM maps indicated model difficulties with approximating certain tissue and background boundaries, secondary to visually imperceptible signal differences, i.e., noise, in these areas. However, as the goal of this work is to improve clinician workflows, these perceived differences are likely not clinically significant. Moreover, these issues have been echoed in similar work (31). Consistent with clinician preferences, our DL model generally had difficulties in successfully synthesizing pathologic tissue, i.e., primary and nodal tumor volumes. This is likely secondary to the relative lack of representation of pathologic tissue in the training set since models are trained at a slice-by-slice level. Our models were trained using patients from a variety of HNC subsites (i.e., oropharynx, nasopharynx, glandular, etc.), so there was substantial heterogeneity in nodal appearance and location, which likely made model training difficult for these subregions. However, while the contrast in these nodal regions was often visibly different in synthetic images, their relative size, shape, and texture may remain similar to the ground-truth image. It has been suggested that geometrical properties, i.e., size, shape, and texture, are often particularly important for image segmentation (58,59). Therefore, cases where tumor volume synthesis “fails” may not necessarily render these images clinically unusable, but additional research should be performed to verify these claims.

Our study is not without limitations. Although our total number of unique image sets was larger than most in the existing HNC synthetic imaging literature (16,18,20,21,24,31), our model training and evaluation was limited to a small cohort from the same institution. However, since model training occurred on a slice-by-slice basis, we utilized on the order of ∼20,000 training data points, which allowed us to leverage DL approaches effectively. Moreover, we only tested one DL approach; several architectural modifications have been proposed that could improve our models in terms of similarity metric performance (31). However, since the end goal of our study is clinician acceptability in an RT workflow and our model passed the Turing test, additional improvement of model performance may be moot. A further limitation of our study is that we used a previously trained DL OAR auto-segmentation model that was developed for 2-minute T2w scans, which limited its generalizability to 6-minute T2w scans. However, the performance of the model on the 6-minute scans was above the expected clinical interobserver variability for several OARs and we were therefore confident in its use for the analysis; interestingly, these results offer evidence for the generalizability of MRI auto-segmentation models, but future studies should further investigate how to optimally translate models developed for different sequences. Additionally, it warrants mentioning that the raw outputs of the DL model were often blurrier than their ground-truth counterparts. This effect has been widely documented in synthetic image studies (15,20,22,24,25,30), often driven by a lower spatial resolution input image up-sampled to match a higher spatial resolution output image. While this slight blurring effect is unlikely to affect underlying image quality (as evidenced by metric performance), in supplementary analyses we demonstrated that this difference was perceived by the clinicians and added bias to the analysis. Therefore, we applied a sharpening filter to synthetic images for the Turing test to remove bias associated with blurry DL outputs, which yielded improved results. While the application of a sharpening filter serves as an effective mitigation technique, as shown in the supplementary analysis, it slightly decreased similarity metric performance. Therefore more sophisticated approaches to decrease image blurriness without the cost of image quality should be investigated, such as the use of super-resolution DL networks (60). Finally, the role of imaging biomarkers, i.e., radiomics (61), is predicted to play an increasing role in MRI-guided ART (3,4). While we have provided minor supplementary analyses on this issue, future work should investigate the feasibility of using synthetic images for radiomic-related analyses.

## Conclusions

In summary, using 2-minute MRI inputs, we designed a CycleGAN DL model to generate synthetic scans that were similar to ground-truth 6-minute scans. As evidenced in quantitative and qualitative analysis, our synthetic scans are of comparable quality to ground-truth 6-minute scans. This model could be used to generate high-quality scans at a reduced acquisition time, thereby improving patient comfort and scanner availability in an MRI-guided ART workflow. Future studies should include external validation of our model, DL architectural improvements, and investigating synthetic images in the context of imaging biomarkers. Moreover, while we study the generation of high-quality output images from standard 2-minute input images, potentially these methods could be extrapolated to input images acquired over an even shorter period (e.g., < 1-minute) to further improve time-savings.

## Data Availability

All DL inputs and outputs for the test set cases are made publicly available on Figshare: 10.6084/m9.figshare.20099252 (private until manuscript acceptance).

## Acknowledgments

We thank Ms. Ann Sutton from the Editing Services Group at The University of Texas MD Anderson Cancer Center Research Medical Library for editing this article. The authors also acknowledge the following individuals for their contributions to the NIH-funded academic-industrial partnership grant (R01DE028290) that funded this work and for their general support and feedback regarding this project: Spencer Marshall, Hafid Akhiat, Michel Moreau, Edyta Bubula-Rehm, Chunhua Men, and Etienne Lessard of Elekta and Alex Dresner of Philips.

