## Appendices for "Deep-Learning-Based Generation of Synthetic High-Resolution MRI from Low-Resolution MRI for Use in Head and Neck Cancer Adaptive Radiotherapy"

### Appendix A. Clinical Data

**Table A1.** Patient demographic characteristics. CUP = cancer of unknown primary.

| Characteristic | Train (n=35) | Test (n=18) |
|---|---|---|
| Age (median, range) | 63 (22-81) | 62 (48-83) |
| Sex | | |
| Male | 28 | 17 |
| Female | 7 | 1 |
| Race | | |
| White | 34 | 15 |
| Other | 1 | 3 |
| Tumor site | | |
| Oropharynx | 18 | 8 |
| Oral | 3 | 2 |
| Nasopharynx | 3 | 1 |
| Larynx | 5 | 3 |
| Node (CUP) | 1 | 3 |
| Volunteer | 3 | 0 |
| Gland | 1 | 1 |
| Other | 1 | 0 |
| T stage | | |
| Tx, 0, or NA | 5 | 5 |
| 1 | 8 | 4 |
| 2 | 12 | 6 |
| 3 | 3 | 0 |
| 4 | 7 | 3 |
| N stage | | |
| Nx, 0, or NA | 16 | 4 |
| 1 | 9 | 6 |

| | | |
|---|---|---|
| 2 | 9 | 5 |
| 3 | 1 | 3 |
| M stage | | |
| 0 or NA | 33 | 15 |
| 1 | 2 | 3 |

**Appendix B. 6-minute MRI vs. 2-minute MRI initial survey**

As initial motivation towards developing a synthetic MRI deep learning model, we created a survey to gauge physician preferences for ground-truth 2-minute vs. ground-truth 6-minute scans. Neuroimaging Informatics Technology Initiative (NIfTI) formatted ground-truth 2-minute scans and 6-minute scans of the 18 test cases were randomly relabeled as either “Image A” or “Image B”. These blinded images were provided to four physician observers to be visualized in 3D Slicer (1). Observers were free to alter the window width and level at their discretion. Observers were instructed to document their preference (“Image A” or “Image B”) in a spreadsheet for a set of regions of interest (ROI)s: right parotid gland, left parotid gland, left submandibular gland, right submandibular gland, spinal cord, brainstem, mandible, primary tumor, and metastatic lymph node(s). Not all images had all ROIs present (i.e., some patients had gland or tumor resections). Observer preferences were remapped to the original image identifiers to determine which observers preferred 6-minute scans vs. 2-minute scans for each ROI (**Figure B1**). With the exception of one observer, who preferred glandular structures on 2-minute scans, all observers overwhelmingly preferred 6-minute scans over 2-minute scans for all ROIs.

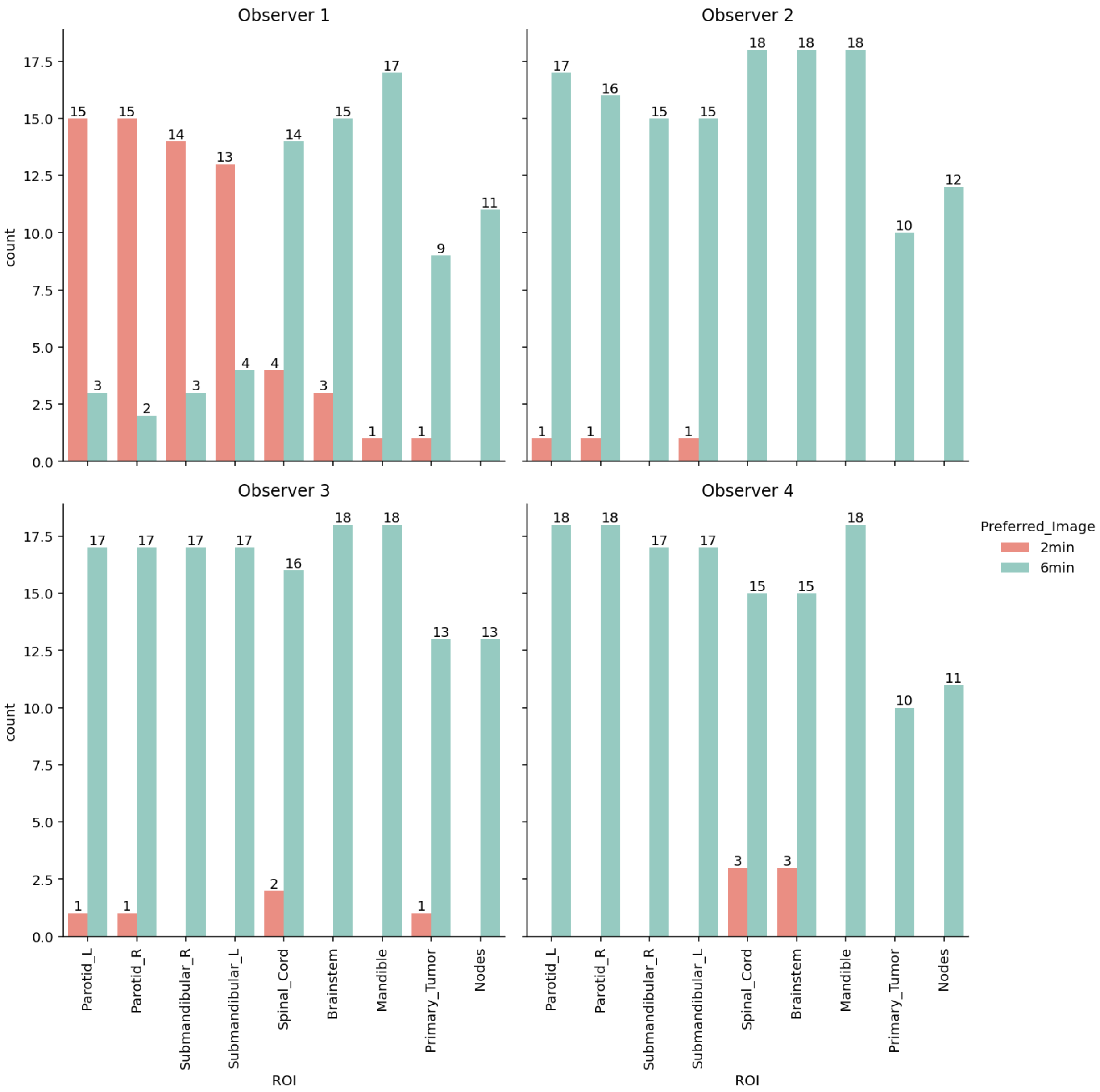


**Figure B1.** Observer preferences for visualizing regions of interest (ROI) on ground-truth 2-minute scans (red) vs. ground-truth 6-minute scans (green) for a variety of regions of interest.

### Appendix C. Additional Auto-segmentation Data

A previously trained head and neck cancer organ at risk (OAR) auto-segmentation model initially developed in independent 2-minute MRI scans was applied to the ground-truth 2-minute, ground-truth 6-minute, and synthetic 6-minute MRI scans in the test set. Examples of auto-segmented OARs overlaid on images and in 3D volumetric format for ground-truth 2-minute and ground-truth 6-minute scans in one case are shown in **Figure C1**.

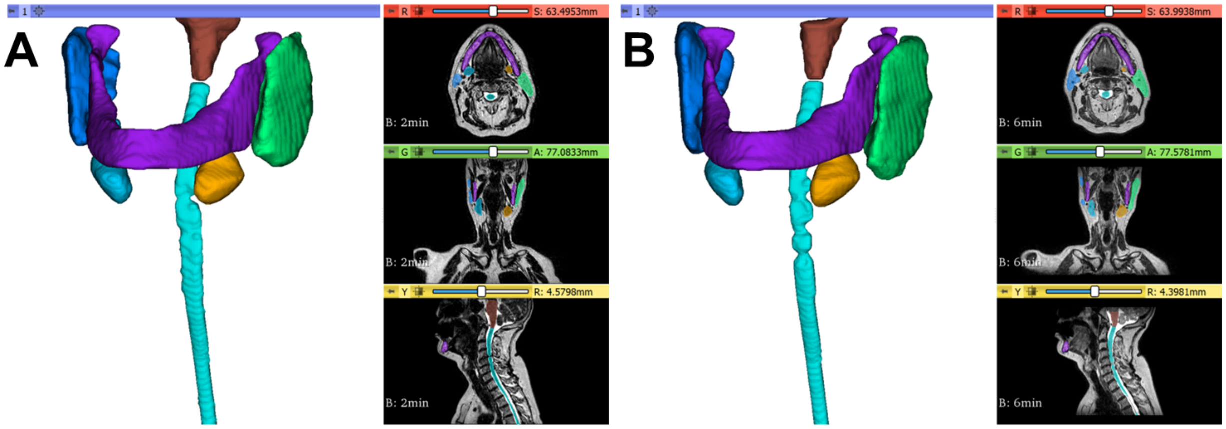


**Figure C1.** Organ at risk auto-segmentation 3D representation and axial/coronal/sagittal views for ground-truth 2-minute **(A)** and ground-truth 6-minute **(B)** scans for 1 representative case where all structures were correctly contoured. Right parotid, left parotid, left submandibular gland, right submandibular gland, spinal cord, brainstem, and mandible, are represented by the dark blue, green, light blue, orange, teal, brown, and purple structures. Visualizations generated in 3D Slicer.

Interobserver variability (IOV) cutoffs for each OAR were determined for the Dice similarity coefficient (DSC) and average surface distance (ASD) from supporting data in work by McDonald et al. (2). For equivalence tests, we also implemented the interquartile range (IQR) values as the minimum (-IQR) and maximum (+IQR) equivalence bounds. A table of the estimated values is shown below (**Table C1**).

**Table C1.** Median (med) and interquartile range (IQR) values for expert interobserver Dice similarity coefficient (DSC) and average surface distance (ASD) for each organ at risk structure.

| Structure | Med DSC | IQR DSC | Med ASD | IQR ASD |
|---|---|---|---|---|
| Left Parotid Gland | 0.83 | 0.08 | 1.80 | 1.05 |
| Right Parotid Gland | 0.84 | 0.07 | 1.50 | 1.00 |
| Mandible | 0.71 | 0.10 | 1.40 | 1.10 |
| Spinal Cord | 0.85 | 0.10 | 0.60 | 0.20 |

| | | | | |
|---|---|---|---|---|
| Brainstem | 0.86 | 0.07 | 1.30 | 0.40 |
| Left Submandibular Gland | 0.75 | 0.12 | 1.40 | 1.20 |
| Right Submandibular Gland | 0.78 | 0.11 | 1.40 | 1.00 |

In the main manuscript we do not include the OARs whose median values between ground-truth 6-minute and ground-truth 2-minute scans do not cross the corresponding IOV median value (i.e., lower than threshold in case of DSC or higher than threshold in case of ASD) as these structures would likely not be clinically acceptable, i.e., spinal cord, brainstem, left/right submandibular glands. However, for completeness, we show the full bar plot representations for all OARs below (**Figure C2**). DSC and ASD equivalence tests (two one-sided t-tests) were non-significant ($p > 0.05$) for the spinal cord, brainstem, left submandibular gland, and right submandibular gland.

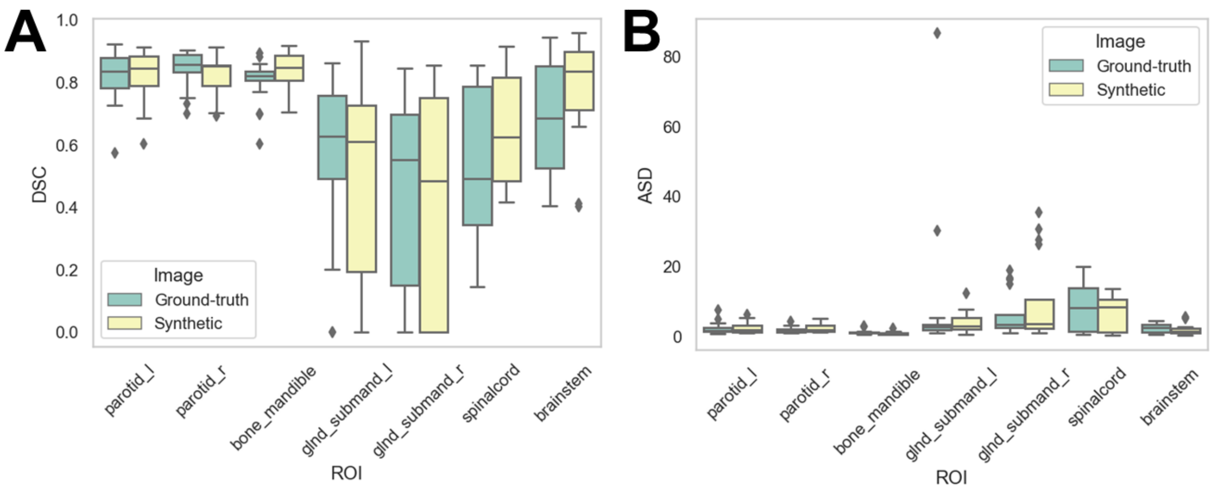


**Figure C2.** Complete auto-segmentation results. Auto-segmented organs at risk were generated on ground-truth (green) or synthetic (yellow) 6-minute images and compared against ground-truth 2-minute images using the **(A)** Dice similarity coefficient (DSC) and **(B)** average surface distance (ASD).

### Appendix D. Additional Image Similarity Data

In **Table D1** we show image similarity metrics (mean squared error [MSE], structural similarity index [SSIM], and peak signal to noise ratio [PSNR]) for the deep learning model without N4 bias field correction, with N4 bias field correction before application of a sharpening kernel (main results described in manuscript), and with N4 bias field correction after application of a sharpening kernel. Generally, metrics improved slightly or remained similar with N4 bias field correction, and worsened after application of the sharpening kernel.

**Table D1.** Image similarity results across the whole image and various subregions for the deep learning model without N4 bias field correction, with N4 bias field correction, and with N4 bias field correction after application of a sharpening kernel. MSE = mean squared error, SSIM = structural similarity index, PSNR = peak signal to noise ratio.

| ROI | Model | MSE | SSIM | PSNR |
|---|---|---|---|---|
| whole | without N4 | 0.19 (0.06) | 0.93 (0.03) | 33.41 (2.13) |
| whole | with N4 | 0.19 (0.05) | 0.93 (0.03) | 33.14 (2.30) |
| whole | with N4 sharpened | 0.22 (0.06) | 0.92 (0.04) | 32.27 (2.17) |
| bone_mandible | without N4 | 1.15 (0.68) | 0.56 (0.09) | 23.38 (2.41) |
| bone_mandible | with N4 | 1.10 (0.47) | 0.56 (0.09) | 23.51 (1.82) |
| bone_mandible | with N4 sharpened | 1.31 (0.57) | 0.54 (0.08) | 22.76 (1.85) |
| brainstem | without N4 | 5.13 (3.12) | 0.67 (0.10) | 19.53 (2.15) |
| brainstem | with N4 | 4.79 (2.96) | 0.66 (0.09) | 19.28 (2.14) |
| brainstem | with N4 sharpened | 5.71 (2.96) | 0.64 (0.09) | 18.63 (1.99) |
| external | without N4 | 0.41 (0.14) | 0.80 (0.05) | 30.25 (1.26) |
| external | with N4 | 0.40 (0.11) | 0.81 (0.05) | 30.23 (1.30) |
| external | with N4 sharpened | 0.46 (0.14) | 0.80 (0.06) | 29.51 (1.35) |
| glnd_submand_l | without N4 | 1.72 (1.36) | 0.41 (0.08) | 18.91 (3.04) |
| glnd_submand_l | with N4 | 1.35 (0.76) | 0.44 (0.11) | 19.77 (3.54) |
| glnd_submand_l | with N4 sharpened | 1.68 (0.82) | 0.41 (0.14) | 18.83 (3.59) |
| glnd_submand_r | without N4 | 1.62 (1.40) | 0.48 (0.17) | 17.97 (3.83) |
| glnd_submand_r | with N4 | 1.37 (0.76) | 0.48 (0.14) | 18.28 (3.37) |

| | | | | |
|---|---|---|---|---|
| glnd_submand_r | with N4 sharpened | 1.70 (1.04) | 0.46 (0.15) | 17.46 (3.13) |
| parotid_l | without N4 | 1.35 (0.69) | 0.47 (0.10) | 19.59 (3.57) |
| parotid_l | with N4 | 1.33 (0.49) | 0.46 (0.11) | 19.07 (2.65) |
| parotid_l | with N4 sharpened | 1.67 (0.60) | 0.43 (0.12) | 18.18 (2.56) |
| parotid_r | without N4 | 1.27 (0.93) | 0.53 (0.09) | 19.81 (2.17) |
| parotid_r | with N4 | 1.15 (0.51) | 0.53 (0.11) | 19.59 (1.95) |
| parotid_r | with N4 sharpened | 1.40 (0.60) | 0.51 (0.12) | 18.71 (1.86) |
| spinalcord | without N4 | 1.90 (2.08) | 0.53 (0.09) | 22.60 (2.58) |
| spinalcord | with N4 | 1.86 (1.94) | 0.52 (0.08) | 22.82 (2.43) |
| spinalcord | with N4 sharpened | 2.42 (2.35) | 0.48 (0.09) | 21.74 (2.18) |

In addition to similarity metric analysis, we also performed a preliminary investigation of radiomic features for synthetic vs. ground-truth images. Specifically, we sought to determine the reliability/repeatability of various radiomic feature classes for the previously segmented OARs on the synthetic images generated by the model described in the main manuscript (with N4 bias field correction before application of sharpening kernel). OAR segmentations on 2-minute scans were resampled to the ground-truth 6-minute and synthetic 6-minute scans using a nearest neighbor interpolator. As before, auto-segmented structures for patients which were not present (i.e., glands post-resection) were not included in the analysis. Radiomic feature extractions were performed on z-score normalized images. Using the open-source toolkit, PyRadiomics (3), we extracted the standard default features from first order statistics (firstorder; 19 features), grey level co-occurrence matrix (glcm; 24 features) gray level run length matrix (glrlm; 16 features), neighbouring gray tone difference matrix (ngtdm; 5 features), and gray level dependence matrix (gldm; 14 features) from OARs on ground-truth 6-minute and synthetic 6-minute scans. The default PyRadiomics extraction parameters, e.g., fixed bin width were applied as recommended. Shape features were not extracted since the OAR mask was the same between the scans. We utilized the two-way mixed effects, consistency, single rater/measurement intraclass correlation coefficient (ICC) provided by the pinguoin Python package (4) to calculate ICC values for each feature class/OAR combination. ICC targets were individual patients, raters were the different images (ground-truth, synthetic), and ratings were the OAR radiomic feature values. ICC values less than 0.5 were categorized as non-reliable, while ICC values greater than or equal to 0.5 were categorized as reliable. The ICC results stratified by OAR and radiomic feature category are shown in **Figure D1**. Generally, a greater number of firstorder features were considered reproducible than non-reproducible for most OARs. For gldm and glrlm, a smaller proportion of features were considered reproducible for most OARs. For ngtdm, only a small number of features for the spinal cord and brainstem were considered reproducible. Finally, for glcm and glszm no features were considered reproducible. Future work should investigate the utility of using synthetic images for radiomic feature calculation in MRI-guided adaptive radiotherapy workflows in greater depth.

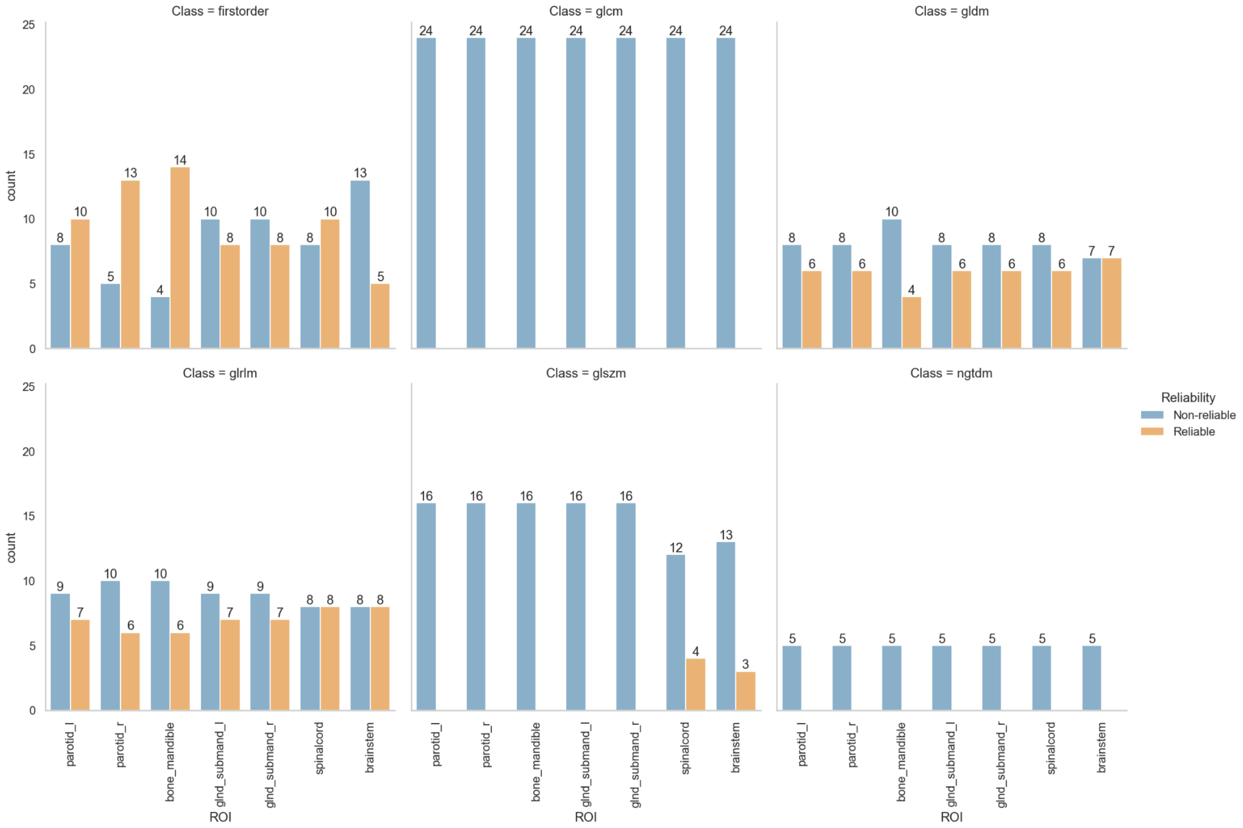


**Figure D1.** Radiomic feature reliability/repeatability on synthetic scans compared to ground-truth scans stratified by region of interest (ROI) and feature category. firstorder = first order statistics, glcm = grey level co-occurrence matrix, glrlm = gray level run length matrix, ngtdm = neighbouring gray tone difference matrix, gldm = gray level dependence matrix.

### Appendix E. Turing Test Additional Data

The Turing test was initially performed with raw image outputs and subsequently after the application of a simple sharpening kernel. The same expert physician observers were given re-randomized slice representations of the same cases one week after the initial Turing test. Only the results after application of the sharpening kernel are displayed in the main manuscript. For completeness we display the results of the Turing test without the sharpening kernel below. As opposed to the results with the application of the sharpening kernel, the original outputs were often distinguishable due to a slight systematic blurring effect. **Table E1** shows the Turing test and clinician preference results while **Figure E1** shows the stratified clinician preference results.

**Table E1.** Turing test and image preference results for three physician expert observers before application of sharpening kernel. Each observer was asked to determine the image identity of blinded paired ground truth (GT) or synthetic 6-minute MRI scan slices in a randomized fashion and also provide their preference. Two one sided tests for two proportions were applied to determine if observer estimates were equivalent to chance.

| Observer | % Correct | p-val | % GT Preference | p-val |
|---|---|---|---|---|
| 1 | 98 | 0.99999997 | 93 | 0.999979105 |
| 2 | 68 | 0.38496071 | 62 | 0.125479502 |
| 3 | 65 | 0.23466347 | 58 | 0.043816053 |

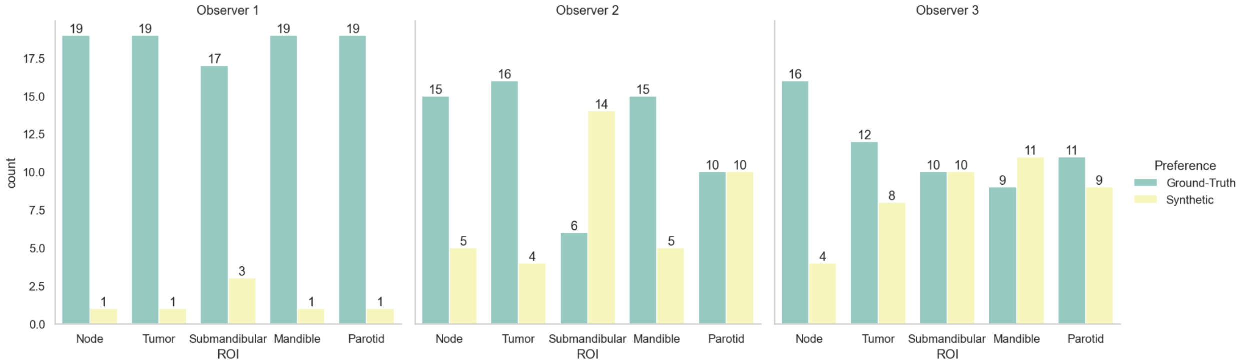


**Figure E1.** Clinician image preferences stratified by region for Turing test before application of sharpening kernel. Green = ground-truth 6-minute MRI slice, yellow = synthetic 6-minute MRI slice.

Additionally, observers were instructed to provide comments where desired to indicate specific reasons for categorizing images as either ground-truth or synthetic. The raw comments for the Turing test before (**Table E2**) and after (**Table E3**) application of the sharpening kernel are shown below for each observer.

**Table E2**. Turing test comments for each observer before application of sharpening kernel.

| Observer | Slice ID | Left image impression | Right image impression | I prefer | Comments |
|---|---|---|---|---|---|
| 1 | 1 | AI | Real | Left | Left has better margins between structures |
| 1 | 2 | Real | AI | Right | Right has better resolution |
| 1 | 5 | Real | AI | Right | Right has better visualization of tissues |
| 1 | 6 | AI | Real | Left | Left has better resolution |
| 1 | 7 | AI | Real | Left | Left has less noise |
| 1 | 9 | AI | Real | Left | Left has better contrast between tumor and surrounding normal tissues |
| 1 | 12 | Real | AI | Right | Right shows better visualization of tumor |
| 1 | 15 | Real | AI | Right | There is better discrimination of tumor in the rigth |
| 1 | 17 | Real | AI | Right | Right shows better discrimination between tumor and submandibular gland |
| 1 | 18 | Real | AI | Right | Right has better visualization of LN |
| 1 | 20 | AI | Real | Left | Left has better margins between tissues |
| 1 | 21 | Real | AI | Right | Right has better discrimination between LN and Rt submandibular gland |
| 1 | 22 | AI | Real | Left | Left has better visualization of tumor |
| 1 | 25 | AI | Real | Left | Left has better quality in defining Lt RP LN |
| 1 | 26 | AI | Real | Left | Left has better visualization of parotids |
| 1 | 27 | Real | AI | Right | Right has better visualization of Rt level IB LN |
| 1 | 28 | Real | AI | Right | Right has better discrimination of Rt cervical LN |
| 1 | 31 | AI | Real | Left | Left has better visualization of tumor |
| 1 | 32 | AI | Real | Left | Left has better visualization of tumor |
| 1 | 34 | AI | Real | Left | Left has better visualization of LN margins |
| 1 | 36 | AI | Real | Left | Left has better visualization of LN margins |
| 1 | 38 | Real | AI | Right | Right has better visualization of normal tissues' margins |
| 1 | 39 | AI | Real | Left | Left has better visualization of normal tissues' margins |

| | | | | | |
|---|---|---|---|---|---|
| 1 | 41 | AI | Real | Left | Left has better visualization of tumor |
| 1 | 44 | AI | Real | Left | Left has better discrimination of Lt RP LN |
| 1 | 45 | AI | Real | Left | Left has better visualization of LN margins |
| 1 | 48 | AI | Real | Left | Left has better demarcation of normal tissues |
| 1 | 53 | AI | Real | Left | Left has better descrimination of primary tumor |
| 1 | 54 | Real | AI | Right | Right has better discrimination of primary and nodal volumes |
| 1 | 57 | Real | AI | Right | Right has better demarcation of LN |
| 1 | 59 | AI | Real | Left | Left has better discrimination of tumor |
| 1 | 63 | AI | Real | Left | Left has better visualization of GTVP, GTVN |
| 1 | 64 | AI | Real | Left | Left has better defining og LN margins |
| 1 | 65 | AI | Real | Left | Left has better discrimination between Lt submandibular gland and LN |
| 1 | 66 | AI | Real | Left | Left has better visualization of Lt RP LN |
| 1 | 71 | Real | AI | Right | LN can be better visualized at right image |
| 1 | 75 | AI | Real | Right | Right has better discrimination of primary and nodal volumes |
| 1 | 79 | Real | AI | Right | Right has better visualization of Rt LN |
| 1 | 80 | Real | AI | Right | Right has better discrimination between LN and Lt submandibular gland |
| 1 | 83 | AI | Real | Left | Left has better discrimination of Lt primary tumor |
| 1 | 84 | AI | Real | Right | Right has better visualization of normal structures |
| 1 | 86 | Real | AI | Right | Better visualization of tumor at right image |
| 1 | 88 | Real | AI | Right | Better detection of Lt RP LN at right image |
| 1 | 89 | Real | AI | Right | Better detection of GTVP, GTVN at right image |
| 1 | 93 | Real | AI | Right | Better discrimination between LN and Rt submandibular gland at right image |
| 1 | 94 | Real | AI | Right | Can't actually see great difference between both |
| 1 | 95 | Real | AI | Left | Can see more noise at Rt image |
| 1 | 100 | Real | AI | Right | Better visualization of LN at right image |
| 2 | 1 | AI | Real | Left | Left has better margins between structures |

| | | | | | |
|---|---|---|---|---|---|
| 2 | 3 | Real | AI | Left | Submandibular gland on Rt looks calccified when it might not |
| 2 | 4 | AI | Real | Right | sublingual glands aren't visible on Lt |
| 2 | 8 | Real | AI | Right | less noise |
| 2 | 16 | Real | AI | Left | sublingual glands not clear |
| 3 | 1 | AI | Real | Right | better details |
| 3 | 6 | AI | Real | Right | Rt vascular vessles seen better |
| 3 | 7 | AI | Real | Right | Rt tumor boundries seen better |
| 3 | 8 | AI | Real | Right | Rt vascular space more detailed |
| 3 | 9 | AI | Real | Left | better LN margins |
| 3 | 10 | AI | Real | Right | vessles better |
| 3 | 11 | Real | AI | Right | tongue mass better delination |
| 3 | 15 | AI | Real | Right | part of Lt vertabrae not clear |
| 3 | 44 | Real | AI | Left | part of image is missing |
| 3 | 46 | AI | Real | Right | hazy vasless and pharynex |
| 3 | 62 | AI | Real | Right | pharyngeal constrictor and vessles betterseen |
| 3 | 66 | AI | Real | Left | better parotid |
| 3 | 68 | Real | AI | Left | tumor better seen |
| 3 | 75 | Real | Real | Right | gross tumor better seen |

**Table E3**. Turing test comments for each observer after application of sharpening kernel.

| Observer | Slice _ID | Left image impression | Right image impression | I prefer | Comments |
|---|---|---|---|---|---|
| 1 | 1 | AI | Real | Left | Left has better margins between structures |
| 1 | 2 | AI | Real | Left | Better visualization of Rt level 2 cervical LN |
| 1 | 3 | Real | AI | Right | Better visualization of tumor at BOT |
| 1 | 6 | AI | Real | Right | Better visualization of submandibular glands at Rt |

| | | | | | |
|---|---|---|---|---|---|
| 1 | 7 | Real | AI | Left | Better demarcation between sbmandibular gland and cervical LN at LT |
| 1 | 8 | Real | AI | Right | Better visualization of Level 2 cervical LN at Rt |
| 1 | 9 | Real | AI | Right | GTV P & N are better seen at Rt |
| 1 | 12 | Real | AI | Right | Better visualization of normal tissues at Rt |
| 1 | 14 | AI | Real | Left | GTVP is better seen at L |
| 1 | 15 | AI | Real | Right | Less noise at Rt |
| 1 | 17 | AI | Real | Right | Better visualization of tumor at Rt |
| 1 | 19 | AI | Real | Left | Level 2 LN is better seen at Lt |
| 1 | 20 | Real | AI | Right | Less noise at Rt |
| 1 | 22 | AI | Real | Left | Better demarcation of submandibular glands at Lt |
| 1 | 23 | AI | Real | Right | Level 2 cervical LN appears cystic at Lt, while it may be not |
| 1 | 24 | AI | Real | Right | Better visulaization of GTVP at Rt |
| 1 | 25 | Real | AI | Left | Better visulaization of GTVP at Lt |
| 1 | 28 | Real | AI | Right | Lt RP is better seen at Rt |
| 1 | 29 | AI | Real | Left | GTVP, GTVN are better seen at Lt |
| 1 | 30 | Real | AI | Left | GTVP, GTVN are better seen at Lt |
| 1 | 35 | Real | AI | Right | Rt has less noise, better visulaization of tumor |
| 1 | 41 | Real | AI | Right | Lt RP is better seen at Rt |
| 1 | 43 | AI | Real | Left | Better visualization of GTVP, GTVN at Lt |
| 1 | 44 | AI | Real | Right | GTVP is better seen at Rt |
| 1 | 45 | AI | Real | Left | GTVN is better seen at Lt |
| 1 | 47 | Real | AI | Left | Tumor is better seen at Lt |
| 1 | 48 | AI | Real | Left | Level 2 cervical LN is better seen at Lt |
| 1 | 50 | AI | Real | Left | GTVP is better seen at Lt |
| 1 | 52 | Real | AI | Left | Less noise at Lt |
| 1 | 55 | AI | Real | Right | Better visualization of tumor |

| | | | | | |
|---|---|---|---|---|---|
| 1 | 56 | AI | Real | Left | Rt level 2 cervical LN is better seen at Lt |
| 1 | 59 | AI | Real | Left | Less noise at Lt |
| 1 | 61 | AI | Real | Left | Lt RP is better seen at Lt |
| 1 | 68 | Real | AI | Left | Tumor is better seen at Lt |
| 1 | 69 | AI | Real | Left | Less noise at Lt |
| 1 | 70 | AI | Real | Right | Tumor is better seen at Rt |
| 1 | 73 | AI | Real | Left | Better demarcation of tissues at Lt |
| 1 | 76 | Real | AI | Right | Tumor is better seen at Rt |
| 1 | 79 | Real | AI | Right | Better denarcation of cervical LN at Rt |
| 1 | 80 | AI | Real | Right | Less noise at Rt |
| 1 | 88 | AI | Real | Left | Better denmarcation of tissues, less noise at Lt |
| 1 | 90 | Real | AI | Left | GTVP is better seen at Lt |
| 1 | 91 | Real | AI | Right | Better demarcation between sbmandibular gland and cervical LN at LT |
| 1 | 94 | Real | AI | Right | Better demarcation of cervical LN at Rt |
| 1 | 96 | Real | AI | Right | Better demarcation of normal tissues |
| 1 | 99 | Real | AI | Right | GTVP is better seen at Rt |
| 1 | 100 | Real | AI | Right | Lt RP is better seen at Rt |
| 3 | 1 | Real | AI | Left | vessles more clear |
| 3 | 2 | AI | Real | Right | artificate on the Rt II |
| 3 | 4 | AI | Real | Right | larynex more clear |
| 3 | 10 | AI | Real | Right | both vertabre not seen |
| 3 | 22 | Real | AI | Left | subman gland more evident |
| 3 | 26 | Real | AI | Left | both tongue is heterogenous |
| 3 | 27 | AI | Real | Right | vertabrae and muscle are bad both |
| 3 | 36 | AI | Real | Right | vertabrae and muscle are bad both |
| 3 | 40 | AI | Real | Right | vertabrae and muscle are bad both |

| | | | | | |
|---|---|---|---|---|---|
| 3 | 54 | Real | AI | Left | not clear details |
| 3 | 65 | AI | Real | Right | disturbed anatomy both in muscles and vetrtabrae |
| 3 | 72 | AI | Real | Left | anatomy not clear vertabrae and muscles |
| 3 | 84 | AI | Real | Right | junction between larynex and vertabrae poorly seen |
| 3 | 93 | AI | Real | Right | disturbed arrangement |
| 3 | 98 | AI | Real | Right | pharnex not clear |
| 3 | 99 | Real | AI | Left | muscles spaces and glands are not clearly lined |
